# Private prescribing of controlled opioids in England, 2014-2021: a retrospective observational study

**DOI:** 10.1101/2023.02.24.23286407

**Authors:** Isabella Martus, Brian MacKenna, William Rial, Jon Hayhurst, Georgia C. Richards

## Abstract

**Background:** Trends of opioid prescribing in the NHS has been well published, yet trends for the same in private prescribing have not been widely established.

**Aim:** To assess trends and geographical variation of controlled opioids prescribed by private prescribers in England.

**Design and setting:** A retrospective observational study in English primary healthcare.

**Methods:** Data on Schedule 2 and 3 controlled opioids (“controlled opioids”) was obtained from the NHS Business Services Authority (BSA) using freedom of information (FOI) requests between 01 January 2014 and 30 November 2021. Absolute counts and rates of the number of items dispensed per cumulative number of registered private prescribers were calculated and stratified over time, by opioid type, and geographical region.

**Results:** 128,341 items controlled opioids were prescribed by private prescribers in England between January 2014 and November 2021, which decreased by 50% from 23,339 items (4.09 items/prescriber) in 2014 to 11,573 items (1.49 items/prescriber) in 2020. Methadone (36%) was the most common controlled opioid prescribed privately, followed by morphine (18%), buprenorphine (16%), and oxycodone (12%). Prescriptions were highest in London (74%), followed by the South-East of England (7%). A proportion of items (n=462, 0.35%) were prescribed by “unidentified doctors” where the prescription is not readily attributable to an individual prescriber by the BSA.

**Conclusions:** Controlled opioids prescribed by private prescribers in England decreased and were primarily prescribed in London. To ensure patient safety, the monitoring and surveillance of controlled opioids dispensed privately should continue and items linked to “unidentified doctors” should be addressed further.

**How this fits in:** There are concerns over the long-term, high-dose use of opioids in people with chronic pain – trends for which have been described using English NHS prescription data. However, opioids can also be acquired from outside of NHS services, including private prescribers, over-the-counter (e.g. co-codamol), and through online healthcare services and pharmacies or the “dark web”. Without exploring non-NHS data, the full picture of opioid use in England cannot be understood. This study sought to fill this gap by investigating opioids in the private sector. We found that the number of controlled opioid items prescribed by private prescribers in England halved between January 2014 and November 2021, and that most prescribing occurred from prescribers in London. There were also controlled opioid items dispensed by “unidentified doctors”, which must be addressed to ensure patient safety. While there is monitoring of controlled drug prescribing by NHS England Controlled Drug Accountable Officers, expanding access to such data to allow for a greater visibility and wider analysis of non-NHS data, including the private prescribing of controlled opioids, will allow policymakers and clinicians to further assess the implementation of pain guidelines and identify harms that can be addressed to improve patient safety.

## Introduction

Opioids are strong analgesics often prescribed in primary care for chronic pain (1–3). There is strong evidence to suggest that the harms of opioids outweigh the benefits when used at high doses and for long durations (4,5). Over the past three decades, opioid prescribing has increased in the UK (2,6), as well as the subsequent increase in opioid dependence, overdose, and deaths associated with their use (7,8).

Studies on the use of opioids in England have focused on prescriptions dispensed in the National Health Service (NHS) or qualitative studies of prescribers in the community (1–3), despite opioids also being available to purchase over-the-counter (e.g. codeine linctus and co-codamol), from online pharmacies or the “dark web”, and through prescriptions from private prescribers (9–11). In an analysis of over-the-counter codeine sales in 31countries, the UK was found to purchase the fourth most products containing codeine (9). Preventable deaths from purchasing medicines online, including opioids, have been reported in England and Wales and an analysis of online marketplaces for controlled substances has found a wide variety and availability of opioids in the UK(10,12). The Care Quality Commission’s (CQC) report on the safer management of controlled drugs briefly mentions trends of some controlled opioids from both NHS and private prescribers, but provides no long term assessment or insights on geographical variation (13–15). A national survey of community pharmacies in 1995 assessed primary and secondary NHS and private methadone prescriptions (11). The findings from this survey described key differences between NHS and private prescribing of opioids, including an increased dispensing of methadone in tablet form (33% private vs. 10.9% NHS) and larger quantity provisions privately rather than daily dispensing in the NHS. The insights from this research would not have been possible if using NHS prescribing alone. Therefore, an up-to-date analysis of opioids dispensed privately is required to better understand the use of opioids in England.

Opioids used in healthcare are controlled under the Misuse of Drugs Act 1971 and the Misuse of Drugs Regulations 2001 as they have the potential to cause harm and are thus subject to increased controls. The legislation aims to prevent the misuse of controlled drugs and the Regulations allocates controlled drugs, including gabapentinoids and opioids into schedules (1 to 5), which set out the controls associated with each schedule (16). These include the specific requirements for private prescriptions for Schedule 2 and 3 controlled drugs. For Schedule 2 and 3 controlled drugs The Shipman Inquiry made several recommendations on the prescribing and monitoring of controlled drugs (17), with one including the introduction of private controlled drug prescriber practitioner codes (18). The Controlled Drugs (Supervision of Management and Use) Regulations 2013 sets out the monitoring requirements. Since 2007, private prescribers in England must write prescriptions for Schedule 2 and 3 controlled drugs on a special prescription form, allowing data to be captured (Box 1)(19). Yet, an analysis has not been openly published to share such data on the recent trends of controlled opioids dispensed by private prescribers. Therefore, the aim of this study was to evaluate trends and geographical variation of controlled opioids prescribed by private prescribers in the English community.

### Box 1

**A summary of how private prescription data for Schedule 2 and 3 controlled drugs are collected from private prescribers by the National Health Service (NHS) Business Services Authorities (BSA)**

- Misuse of Drugs Regulations 2001 mandates private prescriptions of Schedule 2 and 3 controlled drugs (CDs) are written on controlled stationary forms which carries a unique prescriber identification number (PIN).
- Prescribers are assigned a PIN by the NHS BSA following an application process, currently managed by NHS England Controlled Drug Accountable Officers.
- The requisite prescription forms (FP10PCD) are obtained by the prescriber through Primary Care Support England (PCSE).
- Prescription for CD is written by private prescriber with PIN applied.
- Pharmacy dispenses medication and patient signs the back of the prescription form at the pharmacy.
- Forms with unique PIN are submitted to NHS BSA from pharmacy, monthly in arrears.
- Forms scanned by NHS BSA to capture information including prescriber PIN and item prescribed.
- Prescription forms sent to NHS prescription services and included in various internal reports by NHS BSA and included in the EPACT.net but with restricted access (20).

## Methods

### Study design

We designed a retrospective observational study and pre-registered the study protocol on an open repository (21).

### Data sources

We obtained data from the NHS Business Services Authority (BSA) using three freedom of information (FOI) requests to acquire the most up-to-date data (ePACT2, NHS BSA, Copyright 2022. This information is licenced under the terms of the Open Government License)(22–24). Data in ePACT2 is sourced from the NHS BSA Data Warehouse and is derived from products prescribed and dispensed in the community (23). The data summarises the number of controlled opioid prescriptions (“items”), which are Schedule 2 and 3 drugs, dispensed for England at the Area Team Levels (2014-2019) and the Sustainability and Transformation Partnerships (STP) (2020-2021) regions for BNF Chemical Substances under section 7.4.2 in quarterly splits between January 2014 and November 2021.

Data on the number of registered private prescribers in England was obtained from NHS Digital, which had a record of private controlled drug prescribers in England since 2006 (25).

### Data analysis

We combined the three FOI requests and data on the number of registered private prescribers in England in a Google Sheet that was used for analysis. First, we calculated the total crude number of controlled opioid items dispensed between 01 January 2014 and 30 November 2021 and stratified this count by year. For this calculation, we presented a total range as for the second FOI request for 2019 data, NHS BSA did not provide values if the number of items dispensed were below five (24). Thus we estimated a total range using the lowest estimate of one item and the highest estimate of four items. In all figures and other calculations, we used the lowest estimate. We calculated the absolute percentage change overtime between January 2014 and December 2020 because we received 11 months of data for 2021.

To control for changes in the population overtime, we used the number of registered private prescribers as a proxy to calculate the rate. To calulated the rate, we summed the number of registered private prescribers in England from 2006 until 2014 and then cumulatively until 2021. We divided the absolute number of items by this figure to dertemine the rate of controlled opioid items dispensed per registered private prescriber in England. For this calculation, we assumed that all registered private prescriber were active.

To assess types of opioids, we combined BNF Chemical Substances for each chemical name despite salts and combined therpies with opioid antagonists. For example, morphine sulfate was combined with morphine, and buprenorphine HCL/naloxone HCL was combined with buprenorphine. We summed the types of opioids and calculated a percentage of the total for each type and the percentage change over time.

To examine geographical variation, we allocated each area team level (2014-2019) and STPs (2020-2021) to one of the nine regions of England, including London, East of England, East Midlands, West Midlands, North East of England, North West of England, Yorkshire and Humber, South East of England, and South West of England. The number of items of controlled opioids dispensed in each region was summed and declines were used to generate a choropleth map. For the latest data (1 January to 30 November 2021), declines and a choropleth map were used to illustrate the number of items dispensed across STP regions. Percentages were calculated to determine the areas with the greatest and lowest dispensing.

### Software and data sharing

We used Google Sheets to process and analyse the data and to produce line and bar graphs. DataWrapper was used to produce the choropleth map (26). The study protocol, data, and materials are openly available via the Open Science Framework (21,27).

## Results

Between 128,341 and 129,040 controlled opioid items were dispensed by private prescribers in England between January 2014 and November 2021. The volume of controlled opioids prescribed by private prescribers decreased by 50.4% between January 2014 and December 2020 (Figure 1). Controlling for the number of private prescribers in England, trends decreased by 64% from 4.09 items/private prescriber in 2014 to 1.49 items/prescriber in 2020 (Figure 1; Table S1 in Supplement).

**Figure 1:**
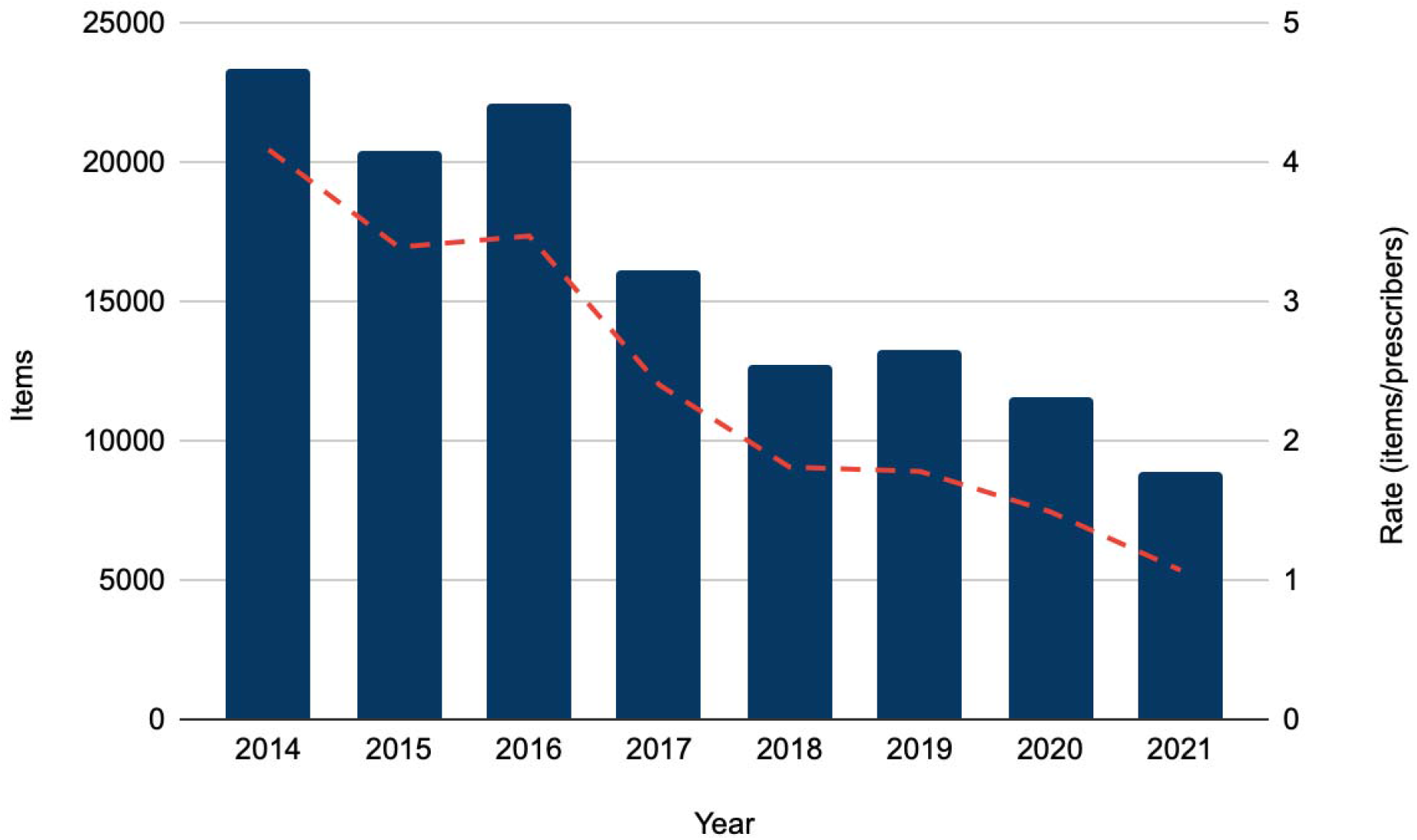
Number of controlled opioid items dispensed by private prescribers in England overtime from 01 January 2014 to 30 November 2021 and the rate of items dispensed by the cumulative number of registered private prescribers.

There were 14 different types of opioids that were dispensed by private prescribers in England (Figure 2). Only codeine preparations for injection are controlled opioids (Schedule 2). Codeine was included by NHS BSA in the first FOI request (2014-2018), but only appeared once in the second quarter of 2018 (22). Methadone was the most common controlled opioid dispensed (36% of total items), followed by morphine (18%), buprenorphine (16%), oxycodone (12%), and tramadol (11%) (Figure 2). Over time, the number of items dispensed decreased for most types of opioids, except for oxycodone and hydromorphone, which increased (Figure 3; Table S2 in Supplement). Five types of opioids, methadone, morphine, buprenorphine, oxycodone and tramadol, represented 93% of all controlled opioids dispensed in England between 2014 and November 2021.

**Figure 2:**
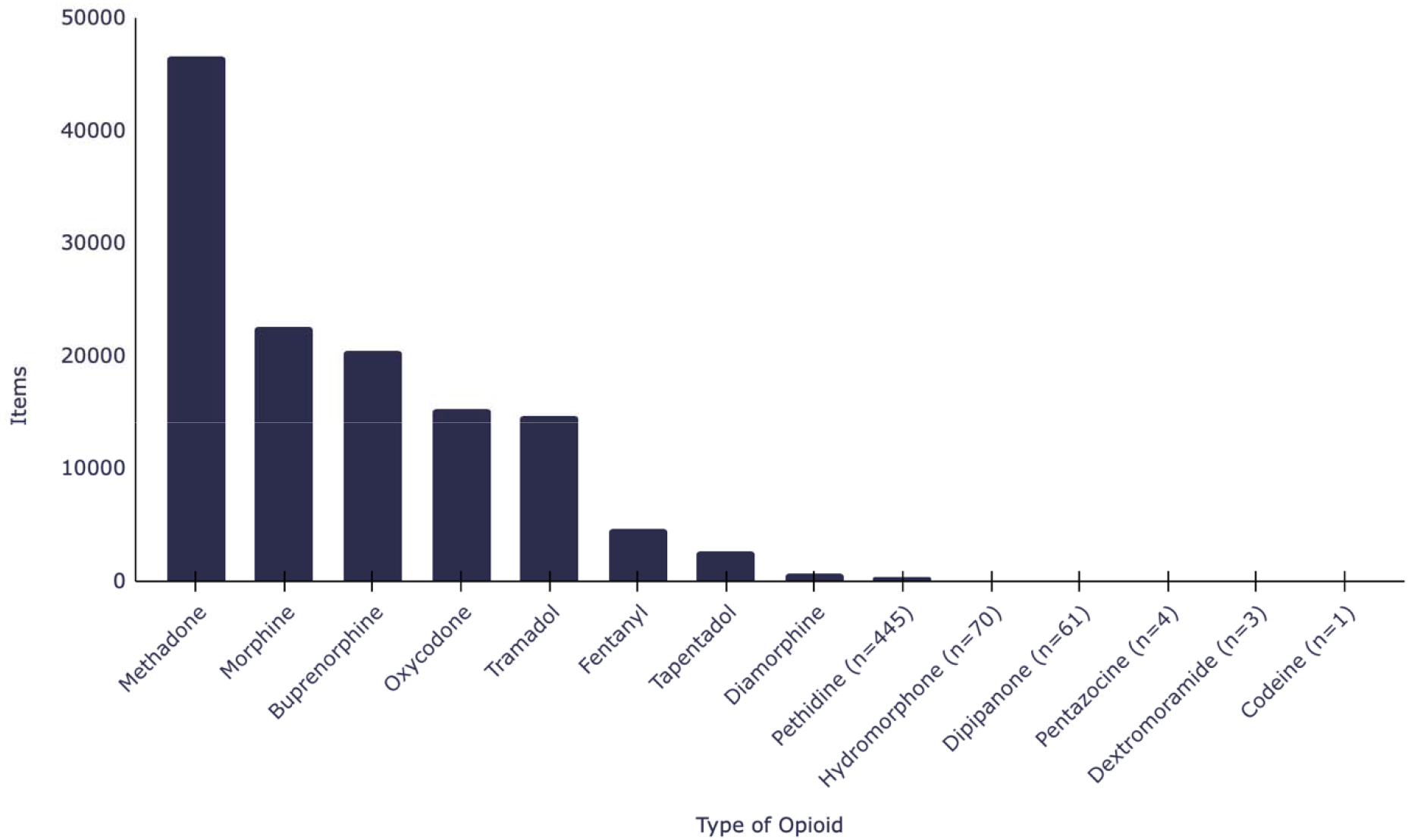
Types of controlled opioids dispensed by registered private prescribers in England between January 2014 and November 2021.

**Figure 3:**
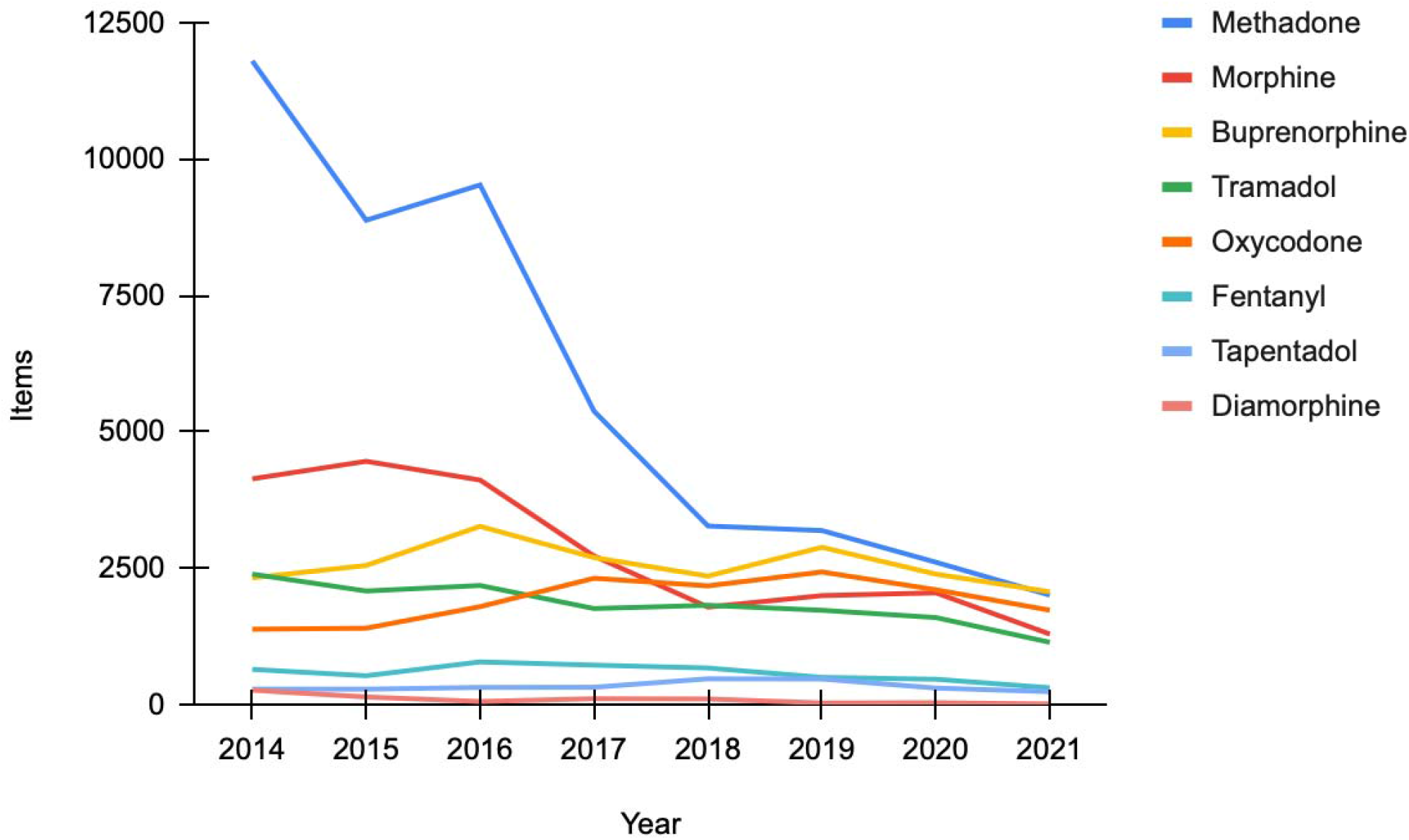
Trends of the top eight most common opioids dispensed by registered private prescribers in England from January 2014 to November 2021.

For the latest data (1 January to 30 November 2021), the North West of London (n=5313) and Staffordshire and Stoke on Trent (n=571) dispensed the most items of opioids, whereas West Yorkshire and Harrogate Health and Care Partnership (n=2) and Somerset (n=2) were in the lowest deciles (Figure 4b; Table S4 in Supplement).

**Figure 4:**
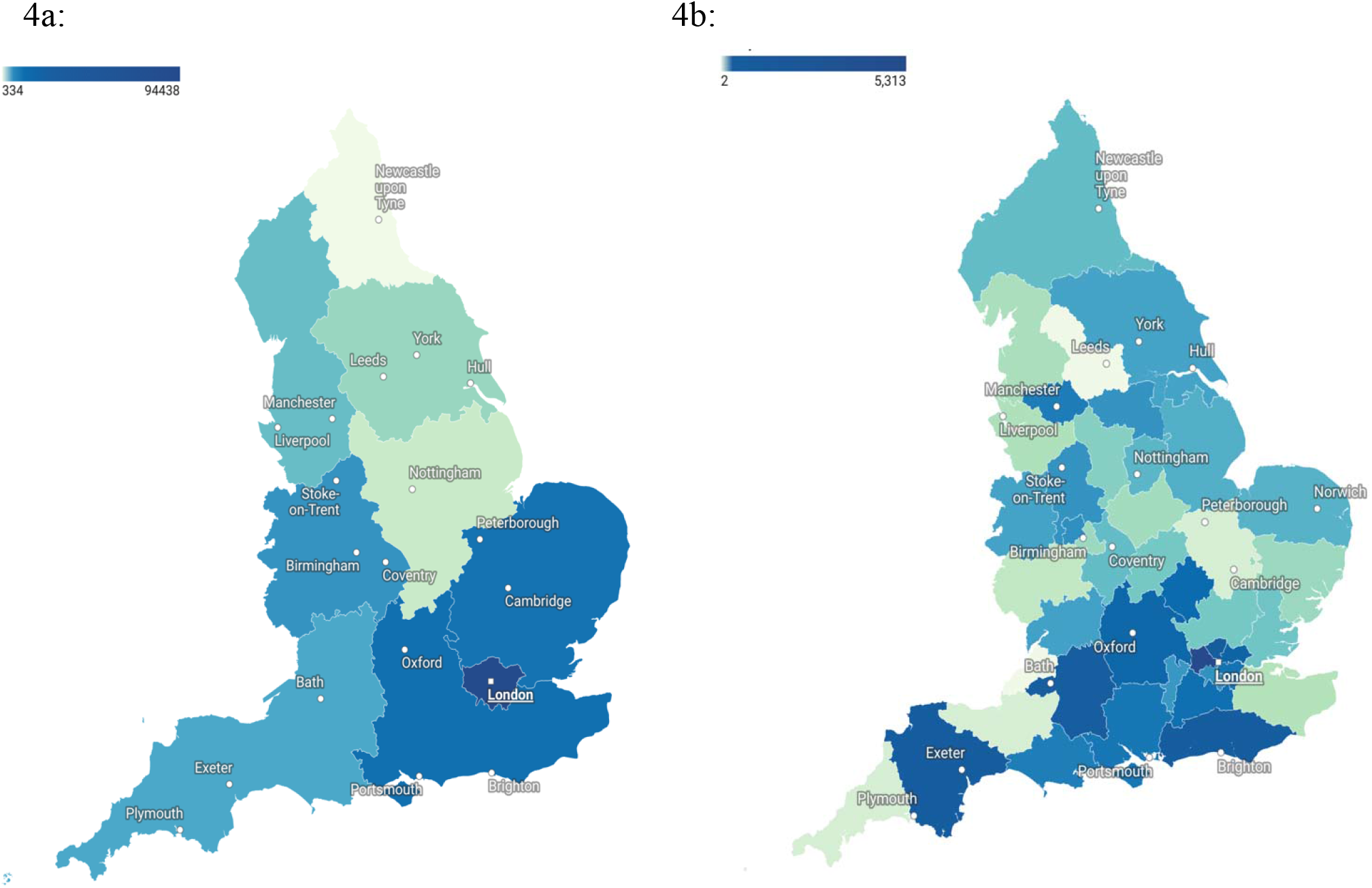
Geographical variation of the absolute number of controlled opioid items dispensed by private prescribers in England, created using declies in Datawrapper. 4a represents the total number of items across the nine regions of England between 1 January 2014 and 30 November 2021. 4b shows the number of items dispensed in each the Sustainability and Transformation Plans (STP) region for the latest data period of 1 January to 30 November 2021. The number of items dispensed from unidentified doctors is not included in these maps.

There were 462 items (0.36%) of controlled opioids that were attributed to “unidentified prescribers” by NHS BSA in 2015 and between January 2018 and November 2021, described as “items that could not be allocated to an individual prescriber” (22–24). Over 53% of these (246 items) were allocated to unidentified doctors in 2021 alone. Morphine (25%), buprenorphine (24%), and methadone (17%) were the most common types of opioids that were attributed to “unidentified prescribers” (Table S5 in Supplement).

## Discussion

Controlled opioids prescribed by private prescribers in England decreased between January 2014 and November 2021. Three-quarters of privately prescribed controlled opioids where from prescribers in London, with methadone and morphine being the most common types of opioids. A small proportion of controlled opioids dispensed privately were attributed to “unidentified prescribers” by NHS BSA.

### Strengths and Limitations

A previous study that examined the use of privately prescribed opioids in England was conducted in 1995 (11), therefore our study provides a much needed update on the trends and geographical variations of controlled opioids dispensed by private prescribers in England. In comparison with previous research that used a sample of data from community pharmacies over one year and focused on injectable methadone for people with opioid addiction (11), our study examined all types of controlled opioids over eight years.

Without access to Electronic Health Records, it is also not possible to assess the doses, duration or indication of use. Instead, the data represented the number of times that a controlled opioid appeared on the prescription form. In the FOI that provided data for 2019, the number of items dispensed were retracted by NHS BSA if the total was less than five. We therefore estimated the the total items using a upper and lower (4 or 1) figure for 2019.

As it is not known what proportion of the English population visited private prescribers over the study period, we standardised the trends by the cumulative number of registered private prescribers in England. Importantly, the data in our study only reflects opioids deemed controlled substances in Schedule 2 and 3 during the study period (January 2014 and November 2021).

Thus our findings do not represent all opioids obtained from private prescribers in England. For example, tramadol was reclassified as a Schedule 3 controlled drug in April 2014, thus we could only receive data for tramadol from this point forward. Opioids in Schedule 4 and 5, which are not subject to the same prescription and monitoring requirements, such as some codeine preparations and oral morphine solution 10mg/5l (Oramorph ®), were not able to be captured in our study. However, the NHS BSA included one item of codeine in the 2014 to 2018 dataset (22).

### Comparison with existing literature

There has been limited information available on the trends of private drug prescribing in England. Opioid research has therefore focused on NHS prescribing (2,6,28), which similarly to our study, found that the prescribing of opioids started to decrease in 2016 (2). The CQC’s annual reports on safer management of controlled drugs simarily found that methadone, fentanyl, morphine, and tramadol decreased between 2020 and 2021 (15,29), and that methadone was the most common private controlled drug in 2014 (30). In our study, oxycodone was the only type of opioid that increased in volume during the study period. A retrospective study that assessed the prescribing of oxycodone in the English NHS between 2013 and 2018 simiarly found an increase in the median rate of oxycodone prescriptions per 1000 population (31). Methadone, which was the most common opioid prescribed by private prescribers in our study is often used for opioid substitution therapy. Thus, this high use in private practice may be driven by the lack of appropriate services and significant regional variation in drug misuse services in England (32).

Previous studies have found geographical variation in the NHS prescribing of opioids across England. In the North of England and in areas of social deprivation, higher rates of opioids have been prescribed (2,8). In contrast, we found that controlled opioids were prescribed more often from London compared with the North of England. The use of private services located in London may be driven by the availability of private clinics and hospitals, the population density, the needs of commuters and visitors to London that require convenient and quick access to healthcare (33), as well as the higher wages which may contribute to the affordability of private doctors in London (34). However, as it is not possible to determine the number of people receiving controlled opioids prescriptions from private prescribers, it is also not possible to establish where they are resident.

### Implications for practice

Examining data on controlled opioids dispensed in the private setting can provide a more comprehensive overview of the total volume of opioids being used in England. Our finding that controlled opioids decreased is in line with the UK’s National Institute for Health and Care Excellence (NICE) guidelines for chronic primary pain in over 16 year olds that advises against the use of opioids (35). However, the 462 items linked to ‘unidentified doctors’ has potential implications for patient safety that should be resolved. Furthermore, codeine, which may not be a Schedule 2 or 3 controlled drug dependiong on the preparation, was included in one of the datasets which should be examined.

Although we were able to access the data through FOI requests, NHS BSA should consider making such data on all controlled drugs publically available without the need for an FOI. If openly available, this data could be integrated into the NHS BSA’s Opioid Comparators (36), and other services such as OpenPrescribing.net who utilize NHS BSA data(37). This would support the analysis of private controlled opioid prescribing over time and allow for the assessment of guidelines and regulations relating to controlled opioids. In the data provided by NHS BSA, the total number of items were redacted if below five for 2019 only. As this was not the case for all other years, NHS BSA should standardised their approach so data is consistent across all requests.

## Conclusion

The private prescribing of controlled opioids in the England has decreased. There was geographical variation with the majority of controlled opioids prescribed privately from London. The data used in our study was obtained using FOI requests, which should be made available and accessible to improve the survellience of controlled opioids. Our findings provide an important insight into another avenue for which people obtain controlled opioids in the community.

## Data Availability

All study materials and data is openly available via online repositories (https://doi.org/10.17605/OSF.IO/HFKDQ(27). The study protocol was preregistered on the Open Science Framework (https://doi.org/10.17605/OSF.IO/HCKPU)(21). Data were obtained from the NHSBSA through freedom of information requests; ePACT2, NHSBSA Copyright 2022. This information is licensed under the terms of the Open Government Licence.

https://doi.org/10.17605/OSF.IO/HFKDQ

## Declarations

## Acknowledgements

The authors would like to acknowledge NHS BSA and NHS Digital who provided us with data for this study.

## Ethical approval

No ethical approval was required for this research.

## Data

All study materials and data is openly available via online repositories (https://doi.org/10.17605/OSF.IO/HFKDQ(27).. The study protocol was preregistered on the Open Science Framework (https://doi.org/10.17605/OSF.IO/HCKPU)(21). Data were obtained from the NHS BSA through freedom of information requests; “ePACT2, NHS BSA Copyright 2022”. This information is licensed under the terms of the Open Government Licence.

## Contributons

GCR submitted the FOI requests and obtained data from NHS digitial. GCR conceptualised, designed, and initiated the study, contributed to data analysis, figure creation, editied the first draft of the manuscript and provided supervisory support and oversight. IEM updated the study protocol, cleaned and analysed the data, produced the figures and tables, wrote the first draft of the manuscript and editied the final draft. BMK provided advice and input into early drafts. WR and JH provided support regarding the controlled drugs legislations and input to manuscript drafts. All study authors read, contributed to, and approved the final manuscript.

