## Supplementary material for "Private prescribing of controlled opioids in England, 2014-2021: a retrospective observational study": S1 in Supplement

**Supplementary materials**

**Table S1**: Number of controlled opioid items, private prescribers, and rate of the number of items dispensed per cumulative number of registered private prescribers in England between 2014 and 2021.

| **Year** | **Total no. of items** | **Cumulative no. of private prescribers** | **Rate (items/prescribers)** |
| --- | --- | --- | --- |
| 2014 | 23339 | 5709 | 4.09 |
| 2015 | 20388 | 6006 | 3.39 |
| 2016 | 22103 | 6361 | 3.47 |
| 2017 | 16075 | 6698 | 2.40 |
| 2018 | 12724 | 7013 | 1.81 |
| 2019^α^ | 13286 | 7450 | 1.78 |
| 2020 | 11573 | 7784 | 1.49 |
| 2021^β^ | 8853 | 8250 | 1.07 |

*^α^In the data from 2019, NHS BSA did not provide absolute values if the number of items dispensed were below five. For this table we present the sum of the lowest estimate of one item for opioids that had less than five items dispensed. ^β^The total number of items in 2021 represent only 11 months of data from January to November.*

**Table S2:** Percentage change in the types of controlled opioids dispensed by registered private prescribers in England between January 2014 and November 2021 in descending order.

| **Type of opioid** | **Number of items** | | **% change** |
| --- | --- | --- | --- |
|  | **2014** | **2021^α^** |  |
| Methadone | 11802 | 2004 | -83% |
| Morphine | 4137 | 1290 | -69% |
| Buprenorphine | 2321 | 2065 | -11% |
| Oxycodone | 1379 | 1728 | 25% |
| Tramadol^β^ | 2390 | 1139 | - 52% |
| Fentanyl | 643 | 305 | -53% |
| Tapentadol | 282 | 234 | -17% |
| Diamorphine | 261 | 9 | -97% |
| Pethidine | 97 | 15 | -85% |
| Hydromorphone | 10 | 15 | 50% |
| Dipipanone | 13 | 7 | -46% |
| Pentazocine | 1 | 0 | n/a |
| Dextromoramide | 3 | 0 | n/a |
| Codeine | 1 | - | n/a |

*^α^The total number of items in 2021 represent only 11 months of data from January to November. ^β^Tramadol became a controlled drug in 2014/2015.*

**Table S3:** Number and percentage of controlled opioid items dispensed by private prescribers in the nine regions of England between January 2014 and November 2021 in ascending order with the colour scale used to develop the deciles in the choropleth map from Figure 4.

| **Region** | **Items** | **% of total items** | **Colour on map (decile)** |
| --- | --- | --- | --- |
| Unidentified doctors | 462 | 0.36 | N/A |
| North East | 334 | 0.26 |  |
| East Midlands | 886 | 0.69 |  |
| Yorkshire and The Humber | 2025 | 1.58 |  |
| North West | 3359 | 2.62 |  |
| South West | 3809 | 2.97 |  |
| West Midlands | 5165 | 4.02 |  |
| Eastern | 8644 | 6.73 |  |
| South East | 9237 | 7.20 |  |
| London | 94438 | 73.58 |  |

**Table S4:** Number of controlled opioid items dispensed across Sustainability Transformation Partnerships (STP) regions in England by private prescribers between January and November 2021.

| **STP regions** | **items** |
| --- | --- |
| Bristol, North Somerset and South Gloucestershire | 2 |
| West Yorkshire and Harrogate | 2 |
| Cambridgeshire and Peterborough | 3 |
| Cornwall and Scilly Isles | 3 |
| Somerset | 3 |
| Herefordshire and Worcestershire | 6 |
| Kent & Medway | 9 |
| Cheshire & Merseyside | 10 |
| Healthier Lancashire & South Cumbria | 11 |
| Leicester, Leicestershire & Rutland | 12 |
| Birmingham and Solihull | 13 |
| Suffolk and North East Essex | 13 |
| Joined Up Care Derbyshire | 16 |
| Hertfordshire and West Essex | 18 |
| Northamptonshire | 18 |
| Mid And South Essex | 19 |
| Coventry and Warwickshire | 21 |
| Cumbria and North East | 21 |
| Nottingham and Nottinghamshire | 28 |
| Norfolk and Waveney | 32 |
| Lincolnshire | 36 |
| Humber, Coast and Vale | 50 |
| Shropshire and Telford and Wrekin | 50 |
| Gloucestershire | 53 |
| Frimley Health & Care | 66 |
| South Yorkshire and Bassetlaw | 70 |
| South West London | 71 |
| The Black Country and West Birmingham | 73 |
| Dorset | 87 |
| Greater Manchester | 88 |
| Hampshire and The Isle Of Wight | 95 |
| Our Healthier South East London | 99 |
| Surrey Heartlands | 105 |
| Bedfordshire, Luton and Milton Keynes | 123 |
| Bucks, Oxfordshire and Berkshire West | 139 |
| East London | 148 |
| Bath and North East Somerset, Swindon and Wiltshire | 175 |
| Devon | 201 |
| Unidentified Doctors | 246 |
| Sussex and East Surrey | 269 |
| North London | 465 |
| Staffordshire and Stoke On Trent | 571 |
| North West London | 5313 |

**Table S5.** Number of controlled opioid items dispensed by unidentified doctors in England by type of opioid in 2015 and between January 2018 and November 2021 in ascending order.

| **Types** | **Items** |
| --- | --- |
| Dipipanone | 1 |
| Pethidine | 3 |
| Tapentadol | 4 |
| Diamorphine | 6 |
| Fentanyl | 14 |
| Tramadol | 63 |
| Oxycodone | 66 |
| Methadone | 79 |
| Buprenorphine | 109 |
| Morphine | 117 |
